# Neutrophil gelatinase-associated lipocalin and Fibrinogen-to-albumin ratio are indicators of nephropathy in adult steady state sickle cell disease patients: A multicenter case-control study in Ghana

**DOI:** 10.1101/2024.10.14.24315440

**Authors:** Stephen Twumasi, Enoch Odame Anto, Christian Obirikorang, Richard Kobina Dadzie Ephraim, Benedict Sackey, Vivian Paintsil, Richard Owusu Ansah, Alfred Effah, Allwell Adofo Ayirebi, Angela Opoku, Godfred Yawson Scott, Leslie Osei, Joyce Duku, Emmanuel Asafo Adjei, Lilian Antwi Boateng

## Abstract

**Background:** Progressive renal failure is one of the main complications in sickle cell disease. Neutrophil Gelatinase-Associated Lipocalin (NGAL) is present in gelatinase/tertiary granules of neutrophils and it is a relatively newly recognized marker of nephropathy. Fibrinogen increases and albumin decreases in inflammatoy conditions like SCD. This study investigated the diagnostic roles of NGAL and fibrinogen-to-albumin ratio in steady state adult sickle cell disease patients.

**Methods:** In this study, 104 sickle cell disease participants and 51 healthy subjects were analysed. Participants’ information was thoroughly documented using a structured questionnaire and patient case records. To evaluate the hematobiochemical parameters, 5ml of venous blood was drawn from each participant and a clean catch of mid-stream urine was collected from each participant. Subjects with sickle cell disease were further classified as SCD nephropathy and SCD non- nephropathy subjects based on reduced estimated Glomerular Filtration Rate (eGFR) and microalbuminuria.

**Results:** Prevalence of nephropathy was 32.7% among adult steady state SCD patients. Significant high levels of Urine Albumin-to-Creatinine Ratio (UACR), NGAL and fibrinogen-to-albumin ratio (FAR) were seen in SCD patients with nephropathy compared to those without nephropathy (*p*<0.001). NGAL levels significantly increased along with increased urine albumin-to-creatinine ratio (r=0.28, *p*<0.005) in both SCD with nephropathy and those without nephropathy. There was a significant negative correlation between creatinine (r=-0.90, *p*<0.0005), urea (r=-0.50, *p*<0.0005) and eGFR in SCD subjects. Similarly, a significant negative relationship existed between UACR and eGFR (r=-0.34, *p*<0.0005). Creatinine also had a significant positive correlation with UACR (r=0.27, *p*<0.005 and urea (r=0.56, *p*<0.0005) in SCD subjects. NGAL was found to be a good predictor of nephropathy in steady state SCD patients with AUC=0.742, *p*<0.0001 when compared with creatinine and urea with AUC=0.618, *p*=0.048 and AUC=0.531, *p*=0.647 respectively. However, FAR could not be used as a predictive marker since it had non-significant poor performance (AUC=0.462, *p*=0.536).

**Conclusion:** NGAL as independent marker, is an early predictor of kidney disease as compared to urea and creatinine. Fibrinogen-to-albumin ratio can be used to track nephropathy treatment in steady state sickle cell disease patients since it is elevated in those with sickle cell nephropathy compared to healthy individuals. These two markers can be added to the available array of test in the management of nephropathy among steady SCD patients.

## Introduction

Renal injury is a common consequence of sickle cell disease (SCD) caused by long-term anemia and disrupted circulation through the medullar capillaries (1). Between 5% and 18% of sickle cell disease patients get renal failure (2). Renal failure affects 4–21% of adult SCD patients due to nephropathy in the United States (3). In Ghana, 39.6% of SCD children with nephropathy were found to have CKD based on decreased eGFR (4). According to a study conducted in Ghana by Ephraim *et al.,* (2015), the prevalence of CKD in the overall SCD population was found to be 39.2%, with adults having a prevalence of 68.4% and children having a prevalence of 31.6% (5). The detection of increasing renal impairment using conventional markers such as serum creatinine levels (Cr), or creatinine clearance (Ccr) and urea are frequently deceptive. The serum creatinine, the primary clinical tool for measuring renal function is insensitive for the detection of moderate alterations in renal function and is affected by factors unrelated to renal activity, such as age, gender, race, and lean muscle mass (6). The equations to estimate glomerular filtration rate do not present a good correlation with laboratory measurements (7). The conventional renal function tests such as serum creatinine, urea and glomerular filtration rate show abnormal results in SCD only when there is extensive renal damage (8). There is a growing need to develop more sensitive markers for the early detection of renal dysfunction in SCD patients since derangement of the conventional markers would be late indicators of irreversible kidney damage.

Neutrophil Gelatinase-Associated Lipocalin (NGAL) is present in tertiary granules of activated neutrophils. Kidney tubular cells may produce NGAL in response to various injuries. It is a relatively newly recognized marker of nephropathy. NGAL was suggested to be a biomarker of acute kidney injury (AKI) even in the setting of chronic kidney disease and has a strong association with albuminuria(9). Following ischemia and acute kidney injury, NGAL is one of the earliest substances that are released into the urine. However, there are limited studies of NGAL in SCD as a marker of renal tubular damage and SCN(10).In cancer studies, the fibrinogen-to-albumin ratio (FAR) was a more significant predictive factor than each indicator itself (11). The purpose of this study is to see if NGAL and FAR could be used as indicators of renal impairment in sickle cell disease patients.

## Materials and Methods

### Study design, duration and study setting

The present study utilized a prospective case-control design and it was conducted at Komfo Anokye Teaching Hospital (KATH), Kumasi South Government Hospital and Suntreso Government Hospital from November 25, 2023 to March 31, 2024. The KATH is located at Bantama, Kumasi, Ashanti Region, Ghana. It is the sole tertiary healthcare facility in the Ashanti Region and the second-largest hospital in Ghana. Kumasi South Hospital is located in Atonsu Agogo and Suntreso Government Hospital is located in Suntreso. These facilities are situated in the Kumasi Metropolis in the Ashanti Region of Ghana. They are a 24- hour working health facilities that boast of services like laboratory, radiology, obstetrics and gynaecology, surgery, paediatrics, internal medicine and emergency among others. They also have a sickle cell clinic from where SCD participants were recruited for the study.

### Ethical Clearance and Informed Consent

The Komfo Anokye Teaching Hospital’s (KATH IRB/AP/175/23) and Kwame Nkrumah University of Science and Technology’s (CHRPE/AP/1037/23) committees on human research and publication ethics approved the study. Once the study participants had received a thorough explanation of the study’s objectives, advantages, risks, and right to withdraw at any time in both English and the local dialect (primarily Twi), their written consent was requested.

### Study population and selection of participants

According to this study, the participants classified as cases comprised individuals who have been medically diagnosed with SCD. The cases were further categorized into sickle cell nephropathy and sickle cell non-nephropathy subjects based on reduced estimated Glomerular Filtration Rate (eGFR) and microalbuminuria. The individuals classified as controls on the other hand, were characterized by their overall good health and absence of sickle cell disease and nephropathy.

The aim of the study was explained to the study participants. Then, the processes involved in the study, including blood and urine sampling, data collection and laboratory assays were explained to the eligible participants with the help of a clinician.

We then purposively recruited a total of 104 SCD patients and 80 healthy individuals, of which 51 healthy individuals were without nephropathy and were analysed. The SCD patients were registered with the sickle cell clinic at the Komfo Anokye Teaching Hospital and Kumasi South Government Hospital. They reported at the clinic for a review as and when directed by their physician. Owing to their clinical history, physicians in both hospitals assessed the SCD patients and declared them fit for the study before they were recruited. Healthy controls were blood donors from the blood bank units of Suntreso Government hospital and Komfo Anokye Teaching Hospital.

SCD patients who are above eighteen (18) years in a steady state for at least one month, and are without history of diabetes mellitus, arterial hypertension, neoplastic, cardiovascular, renal, lung or endocrine disease were eligible for this study. Also, SCD participants who had symptoms suggestive of sickle cell pain crisis, acute sickness (fever or required referral to an urgent care facility), clinically suspected urinary tract infection and extensive haematuria were excluded. Healthy individuals without history of SCD, diabetes mellitus, arterial hypertension, neoplastic, cardiovascular, inflammatory, renal, lung and endocrine diseases were also eligible for the control group.

### Data collection

Important demographic and clinical history information about the participants were recorded using a well-structured questionnaire and patient case records. Animal protein consumption(fish,meat &egg) were categorised qualitatively as occurring rarely (one per month), irregularly (seven times per month), and regularly (twice per month). A wall-mounted ruler was used to measure height. A Zhongshan Camry Electronic Co. Ltd. bathroom scale (Guangdong, China) was used to assess weight. Estimates were made for height to the nearest centimetre without shoes and weight to the nearest 0.1 kg when wearing light clothes.

### Sampling and analysis

Blood samples were collected following standard protocol (World Health Organization, 2010). Five (5) millilitres (ml) (venous blood) were pulled from each participant and collected into K3 EDTA tube, sodium citrate tube and Serum Separator tube (SST) – 1.8ml into sodium citrate tube first, 2.2 ml separated into SST second and 1ml into 4ml EDTA tube. Whole blood in EDTA was used for Haemogram analysis. Serum in SST was used to analyze liver function markers, creatinine, urea, electrolytes (sodium, potassium and chloride) and NGAL. Sodium citrate was used for fibrinogen measurement. Participants were directed to collect a mid-stream urine in a cleaned dry urine container. Mid-stream urine sample was used for UACR.

### Laboratory assays

Participants’ haemoglobin variants were confirmed by Hb electrophoresis. Full Blood Estimation was done using five (5)- part Automated Haematology Analyser XN-550 (XN-550; Sysmex Corporation, Kobe Japan). Kidney function tests (electrolytes, urea, and creatinine), liver function tests (bilirubin, AST,ALT, ALP, GGT Total Protein and albumin), urine albumin-to-creatinine ratio were determined on Vitros 5600 integrated systems. Estimated GFR (eGFR) was calculated using the Chronic Kidney Disease Epidemiology Collaboration creatinine equation (2009) (eGFR_creat_). Normoalbuminuria and microalbuminuria were defined as urine microalbumin:creatinine ratio < 30 mg/g and ≥ 30mg/g respectively. Fibrinogen was measured with Wonfo Fibrinogen reagent kit and OCG-102 Coagulation analyzer. Fibrinogen-to-Albumin ratio calculations involved dividing the Fibrinogen (FIB) by serum Albumin (ALB) to arrive at values. A commercially available ELISA kit (Melson Shanghai Chemical Ltd, China) was used for the sNGAL measurement. Reagent was used in accordance with the manufacturer’s instructions to measure samples from the controls and the participants using the solid phase ELISA technique. The commonly accepted reference value for serum NGAL is 5.68±1.34µg/L.

### Determination of sickle cell nephropathy

The 2024 KDIGO classification (12) for establishing chronic kidney disease guidelines was used. Reduced eGFR and microalbuminuria were the main determinants of sickle cell nephropathy. For eGFR, nephropathy was classified as individual with reduced eGFR (<60mL/min/1.73m²) and without nephropathy (≥60mL/min/1.73m²). Estimated GFR (eGFR) was calculated using the Chronic Kidney Disease Epidemiology Collaboration creatinine equation (2009) (eGFRcreat) (13). Microalbuminuria was determined by urine albumin-to-creatinine ratio (UACR) greater than or equal to 30mg/g and normoalbuminuria was defined as UACR less than 30 mg/g.

### Statistical analysis

The study’s data were entered and cleaned in Microsoft Excel (2016).The date were exported to R programming version 4.2.3, and IBM Statistical Package for Social Sciences (version 26.0) for analysis. Continuous variables were expressed as mean ± standard deviation or median interquatile ranges depending on the normality while categorical variables were expressed as frequency and percentages. The association of participants baseline parameters between the study groups was investigated using the chi-square test. In addition, the post-hoc Bonferroni test and one-way ANOVA or Kruskal Wallis test followed by pairwise multiple comparisons were used to evaluate the variations in continuous variable between study groups, depending on the normality. To examine the relationship between sNGAL, eGFR, FAR, and UACR, Spearman’s correlation was employed and results represented as heatmaps. Using the Receiver Operator Characteristics Curve, the area under the curves for sNGAL, Urea, Creatinine and FAR for predicting sickle cell nephropathy were determined. All computations of statistical significance were performed using a 95% confidence interval and a *p-value* of 0.05.

## Results

### Baseline characteristics of study participants

A total of 104 patients (46 males and 58 females) with SCD were recruited. Seventy four (74) were HbSS and thirty were (30) were HbSC. They were all in steady state based on their clinical status assessed by haematologists. Fifty one (51) healthy individuals (13 males and 38 females) were analysed. Following the 2024 KDIGO classification (12) for establishing chronic kidney disease guidelines, 34(32.7%) out of the 104 SCD patients had microabuminuria. Subsequently, eleven (11) out of the thirty-four (34) SCD participants with microalbuminuria (UACR≥30) had reduced eGFR (<60mL/min/1.73m²), because microalbuminuria is the superior sensitive marker for kidney damage as indicated in the KDIGO guidelines, 34(32.7%) of SCD particiapnts were diagnosed with nephropathy while 70(67.3%) were without nephropathy. Gender varied significantly across the groups *(p=0.017)* with 96(61.9%) being females and males being 59(38.1%). Again, the distribution of age across the cases and controls was statistically significant *(p=0.013)* as majority of them were less than 25 years 84(54.2%) and were more in the SCD non-nephropathy group 42(50.0%). The participants’ BMI varied significantly among the cases and controls *(p<0.0001)* as majority were having normal weights with least being obese 2(1.3%). Nearly all 152(98.1%) rarely smoked tobacco and the difference across the groups was significant *(p=0.044)* and were rarely 139(89.7%) taking in alcohol *(p=0.004)*. Although, there were no significant relation *(p>0.05)* across the groups but larger proportion of the participants irregularly exercise 101(62.2%), and take in salt 97(62.6%), regularly consume meat & fish 122(78.7%), and egg 126(81.3%) (**Table 1).**

**Table 1:** Baseline characteristics of study participants.

| Variables | Total<br>N=155 | Control<br>n=51 | Cases=104(67.1%) |  | *p*-value |
| --- | --- | --- | --- | --- | --- |
|  |  |  | SCD non-nephropathy<br>n=70 | SCD<br>Nephropathy<br>n=34 |  |
| **Age Group (years)** |  |  |  |  | **0.013** |
| <25 | 84(54.2) | 18(31.4) | 42(50.0) | 24(28.6) |  |
| 25-34 | 38(24.5) | 14(36.8) | 16(42.1) | 8(21.1) |  |
| 35-44 | 19(12.3) | 11(57.9) | 7(36.8) | 1(5.3) |  |
| >44 | 14(9.0) | 8(57.1) | 5(35.7) | 1(7.1) |  |
| **Gender** |  |  |  |  | **0.017** |
| Male | 59(38.1) | 13(22.0) | 35(59.3) | 11(18.6) |  |
| Female | 96(61.9) | 38(39.6) | 35(36.5) | 23(24.0) |  |
| **BMI** |  |  |  |  | **<0.0001** |
| Underweight | 9(5.8) | 0(0.0) | 4(44.4) | 5(55.6) |  |
| Normal | 101(65.2) | 23(22.8) | 55(54.5) | 23(22.8) |  |
| Overweight | 43(27.7) | 28(65.1) | 9(20.9) | 6(14.0) |  |
| Obese | 2(1.3) | 0(0.0) | 2(100.0) | 0(0.0) |  |
| **SCD Phenotype** |  |  |  |  | 0.819* |
| HbSC | 30(28.8) | - | 21(70.0) | 9(30.0) |  |
| HbSS | 74(71.2) | - | 49(66.2) | 25(33.8) |  |
| **Vigorous Exercise** |  |  |  |  | 0.784 |
| Regular | 46(29.7) | 16(34.8) | 22(47.8) | 8(17.4) |  |
| Irregular | 101(65.2) | 33(32.7) | 45(44.6) | 23(22.8) |  |
| Rarely | 8(5.2) | 2(25.0) | 3(37.5) | 3(37.5) |  |
| **Meat&Fish Consumption** |  |  |  |  | 0.508 |
| Regular | 122(78.7) | 38(31.1) | 59(48.4) | 25(20.5) |  |
| Irregular | 31(20.0) | 12(38.7) | 10(32.3) | 9(29.0) |  |
| Rarely | 2(1.3) | 1(50.0) | 1(50.0) | 0(0.0) |  |
| **Egg Consumption** |  |  |  |  | 0.056 |
| Regular | 126(81.3) | 36(28.6) | 61(48.4) | 29(23.0) |  |
| Irregular | 29(18.7) | 15(51.7) | 9(31.0) | 5(17.2) |  |
| **Tobacco Smoking** |  |  |  |  | **0.044** |
| Irregular | 3(1.9) | 3(100.0) | 0(0.0) | 0(0.0) |  |
| Rarely | 152(98.1) | 48(31.6) | 70(46.1) | 34(22.4) |  |
| **Alcohol Intake** |  |  |  |  | **0.004** |
| Regular | 2(1.3) | 2(100.0) | 0(0.0) | 0(0.0) |  |
| Irregular | 14(9.0) | 10(71.4) | 2(14.3) | 2(14.3) |  |
| Rarely | 139(89.7) | 39(28.1) | 68(48.9) | 32(23.0) |  |

| **Salt Intake** |  |  |  |  | 0.066 |
| --- | --- | --- | --- | --- | --- |
| Regular | 47(30.3) | 14(29.8) | 21(44.7) | 12(25.5) |  |
| Irregular | 97(62.6) | 29(29.9) | 47(48.5) | 21(21.6) |  |
| Rarely | 11(7.1) | 8(72.7%) | 2(18.2) | 1(9.1) |  |

BMI: Body Mass Index, HbSS; Haemoglobin SS, HbSC; Haemoglobin SC, Data presented as frequency (%). Chi- square *p*-values were presented, *p*<0.05 was considered statistically significant. Bolded *p*-value was statistically significant, \* ; Fisher Exact test computed for the cases only

### Haemato-biochemical Profile of SCD nephropathy, SCD non-nephropathy and controls

Both diastolic and systolic blood pressures differed significantly across groups. Creatinine was elevated in the SCD nephropathy group (*p*=0.0320), compared to SCD non-nephropathy group. Electrolyte levels such as sodium and potassium were significantly different across groups, with controls showing lower levels of sodium and potassium (*p*<0.0001 and *p*=0.0020, respectively). AST, ALP, and total bilirubin were also significantly different across groups (*p*<0.0001). Moreover, hematological indices including RBC count, hematocrit, and hemoglobin were significantly lower in the SCD nephropathy group (*p*<0.0001), whilst inflammatory markers like WBC, neutrophil count, and monocyte count were higher (*p*<0.0001). Serum NGAL and eGFR levels also showed significant differences, with SCD nephropathy patients having the highest sNGAL (*p*<0.0001) and the lowest eGFR (*p*=0.0250) **(Table 2).**

**Table 2:** Haemato-biochemical Profile of SCD patients nephropathy, SCD non-nephropathy and controls.

| Variable | SCD non-nephropathy<br>(n = 70) | SCD Nephropathy<br>n = (34) | Controls<br>(n = 51) | p-value |
| --- | --- | --- | --- | --- |
| Diastolic Blood Pressure (mm Hg) | 71.00 (67.00 - 76.00) | 71.50 (64.75 - 76.50) | 76.00 (74.00 - 79.00) | <0.0001<sup>b,c</sup> |
| Systolic Blood Pressure (mm Hg) | 119.00 (112.00 - 121.25) | 119.00 (112.00 - 127.00) | 112.00 (109.00 - 119.00) | <0.0001<sup>b,c</sup> |
| Urea (mmol/L) | 2.10 (1.58 - 2.87) | 2.19 (1.31 - 5.05) | 2.60 (1.63 - 3.03) | 0.7890 |
| Creatinine (mmol/L) | 53.50 (44.00 - 76.25) | 72.00 (47.00 - 156.00) | 68.00 (52.00 - 77.00) | **0.0320<sup>a</sup>** |
| BUN/Creatinine ratio | 18.15 (13.48 - 24.90) | 17.35 (13.55 - 23.05) | 16.40 (12.50 - 21.10) | 0.2300 |
| Sodium (mmol/L) | 142.00 (141.00 - 144.00) | 143.00 (140.75 - 145.00) | 140.00 (139.00 - 142.00) | <0.0001<sup>b,c</sup> |
| Potassium (mmol/L) | 4.60 (4.50 - 5.00) | 4.90 (4.60 - 5.20) | 4.40 (4.00 - 4.80) | **0.0020<sup>c</sup>** |
| Chlorine (mmol/L) | 106.00 (102.00 - 109.00) | 107.00 (102.00 - 110.00) | 104.00 (103.00 - 107.00) | 0.2780 |
| Aspartate Transaminase (U/L) | 47.55 (37.38 - 59.13) | 50.75 (38.90 - 70.25) | 32.00 (28.40 - 38.90) | <0.0001<sup>b,c</sup> |
| Alanine Transaminase (U/L) | 23.90 (20.83 - 32.35) | 25.20 (19.65 - 28.65) | 24.80 (17.60 - 31.40) | 0.8070 |
| Alkaline Phosphatase (U/L) | 103.05 (77.83 - 135.55) | 99.20 (71.63 - 166.33) | 72.10 (59.40 - 94.30) | <0.0001<sup>b,c</sup> |
| GGT (U/L) | 41.50 (24.88 - 56.53) | 36.15 (24.55 - 53.25) | 26.80 (22.70 - 40.50) | **0.0140<sup>b</sup>** |
| Albumin (g/L) | 41.80 (39.88 - 43.80) | 41.60 (38.55 - 44.05) | 42.20 (39.30 - 44.60) | 0.4500 |
| Total Bilirubin (mmol/L) | 27.00 (15.78 - 38.88) | 29.55 (15.38 - 41.65) | 6.10 (4.10 - 11.30) | <0.0001<sup>b,c</sup> |
| Direct Bilirubin (mmol/L) | 1.40 (0.00 - 5.10) | 2.65 (0.00 - 5.90) | 1.80 (0.90 - 3.00) | 0.4970 |
| Indirect Bilirubin (mmol/L) | 24.25 (13.50 - 35.05) | 24.25 (13.93 - 39.33) | 4.10 (3.10 - 8.10) | <0.0001<sup>b,c</sup> |
| Fibrinogen (g/L) | 4.31 (3.39 - 5.17) | 4.35 (2.71 - 5.12) | 2.40 (2.01 - 3.24) | <0.0001<sup>b,c</sup> |
| FAR | 0.10 (0.08 - 0.12) | 0.11 (0.07 - 0.12) | 0.06 (0.05 - 0.08) | <0.0001<sup>b,c</sup> |
| RBC (10<sup>6</sup>/mL) | 3.08 (2.52 - 3.94) | 2.73 (2.43 - 3.47) | 4.42 (3.96 - 4.85) | <0.0001<sup>b,c</sup> |
| Hematocrit (%) | 25.75 (22.90 - 29.68) | 24.50 (21.75 - 30.13) | 35.60 (31.90 - 39.50) | <0.0001<sup>b,c</sup> |
| MCV (fL) | 85.80 (74.75 - 96.50) | 88.30 (78.00 - 94.78) | 81.40 (76.40 - 85.10) | **0.0170<sup>b,c</sup>** |
| MCH (pg) | 29.55 (25.98 - 32.78) | 30.30 (27.25 - 32.85) | 27.70 (25.00 - 29.20) | **0.0020<sup>b,c</sup>** |
| MCHC (g/dL) | 35.35 (32.75 - 36.60) | 34.70 (32.85 - 36.03) | 33.90 (32.60 - 34.70) | **0.0060<sup>b</sup>** |
| RDW-SD (fL) | 59.00 (46.85 - 66.30) | 61.90 (50.98 - 71.53) | 40.10 (38.20 - 42.80) | <0.0001<sup>b,c</sup> |
| RDW-CV (%) | 19.20 (17.08 - 22.73) | 19.80 (17.50 - 25.35) | 13.80 (12.90 - 14.60) | <0.0001<sup>b,c</sup> |
| PLT (10<sup>3</sup>/mL) | 279.00 (171.75 - 375.50) | 325.00 (214.25 - 455.25) | 248.00 (205.00 - 292.00) | **0.0490<sup>c</sup>** |
| PDW (fL) | 11.80 (9.90 - 13.50) | 12.45 (10.48 - 14.73) | 12.40 (11.40 - 13.70) | 0.2510 |
| MPV (fL) | 10.50 (9.48 - 11.50) | 11.00 (10.00 - 11.98) | 10.90 (10.40 - 11.40) | 0.1860 |
| PCT (%) | 0.30 (0.20 - 0.40) | 0.36 (0.23 - 0.51) | 0.27 (0.24 - 0.33) | 0.0550 |
| WBC ($10^3$/mL) | 8.38 (6.03 - 10.97) | 8.91 (7.70 - 9.85) | 5.57 (4.90 - 6.85) | **<0.0001<sup>b,c</sup>** |
| Lymphocyte Count ($10^3$/mL) | 3.37 (2.47 - 4.21) | 3.44 (2.89 - 3.89) | 2.18 (1.87 - 2.63) | **<0.0001<sup>b,c</sup>** |
| Monocyte Count ($10^3$/mL) | 0.68 (0.46 - 0.99) | 0.72 (0.46 - 0.93) | 0.53 (0.43 - 0.63) | **0.0050<sup>b,c</sup>** |
| Neutrophil Count ($10^3$/mL) | 3.46 (2.65 - 5.35) | 4.25 (3.32 - 5.23) | 2.82 (1.93 - 3.60) | **<0.0001<sup>b,c</sup>** |
| Eosinophil Count ($10^3$/mL) | 0.13 (0.07 - 0.26) | 0.15 (0.10 - 0.25) | 0.10 (0.05 - 0.16) | **0.0140<sup>b,c</sup>** |
| Basophil Count ($10^3$/mL) | 0.06 (0.03 - 0.10) | 0.06 (0.04 - 0.09) | 0.03 (0.02 - 0.04) | **<0.0001<sup>b,c</sup>** |
| Neutrophil to Lymphocyte Ratio | 1.14 (0.82 - 1.61) | 1.15 (1.03 - 1.56) | 1.25 (0.95 - 1.68) | 0.6270 |
| IG# ($10^3$/mL) | 0.06 (0.03 - 0.09) | 0.05 (0.03 - 0.11) | 0.01 (0.01 - 0.02) | **<0.0001<sup>b,c</sup>** |
| Nucleated RBC ($10^3$/mL) | 0.05 (0.02 - 0.11) | 0.07 (0.04 - 0.25) | 0.00 (0.00 - 0.01) | **<0.0001<sup>b,c</sup>** |
| Reticulocyte Count ($10^6$/mL) | 0.15 (0.12 - 0.22) | 0.17 (0.12 - 0.27) | 0.04 (0.02 - 0.05) | **<0.0001<sup>b,c</sup>** |
| Total Protein (g/L) | 87.38 (83.05 - 92.66) | 87.96 (81.49 - 95.05) | 78.28 (72.75 - 85.01) | **<0.0001<sup>b,c</sup>** |
| Globulin (g/L) | 45.50 (42.20 - 50.48) | 45.40 (41.08 - 54.40) | 36.40 (32.30 - 40.10) | **<0.0001<sup>b,c</sup>** |
| Hemoglobin (g/dL) | 9.00 (8.00 - 10.40) | 8.65 (7.55 - 9.80) | 11.90 (10.80 - 13.20) | **<0.0001<sup>b,c</sup>** |
| Platelet to Lymphocyte Ratio | 78.38 (56.40 - 115.50) | 104.79 (68.17 - 141.26) | 116.35 (85.22 - 151.41) | **<0.0001<sup>b,c</sup>** |
| sNGAL (μg/L) | 5.72 (4.76 - 6.52) | 6.64 (6.23 - 7.29) | 4.47 (3.95 - 4.87) | **<0.0001<sup>a,b,c</sup>** |
| eGFR (mL/min/1.73m<sup>2</sup>) | 139.07 (96.97 - 183.09) | 95.73 (47.80 - 166.43) | 120.66 (106.23 - 155.49) | **0.0250<sup>a</sup>** |

BUN-Blood Urea Nitrogen, WBC- White Cell Count, RBC- Red Blood Count, MCV-Mean Cell Volume, MCH- Mean Cell Haemoglobin, MCHC- Mean Cell Haemoglobin Concentration, PLT- Platelet Count, PDW- Platelet Distribution Width, MPV-Mean Platelet Volume, P-LCR- Platelet- Large Cell Ratio, PCT- Plateletcrit, eGFR-Estimated Glomerular Filtration Rate. <sup>a</sup>: significant when SCD non-nephropathy is compared to SCD nephropathy, <sup>b</sup>: significant when SCD non-nephropathy is compared to Control, <sup>c</sup>: significant when SCD nephropathy is compared to Control. Kruskal Wallis p-values were presented, $p < 0.05$ and bolded means statistically significant

### Correlation between sNGAL and biochemical parameters

There was a significant positive correlation between sNGAL and UACR, a significant negative correlation between UACR and eGFR. There was a weak but significant correlation between potassium and sNGAL (r=0.29, *p*<0.005), a significant moderate negative corelation between eGFR and urea (r= -0.50, *p*<0.0005) and a strong negative correlation with creatinine as it is already established (r=-0.90, *p*<0.0005). UACR also correlated weakly but significantly with creatinine in a direct proportion (r=0.27, *p*<0.005) (**Figure 1).**

**Figure 1:**
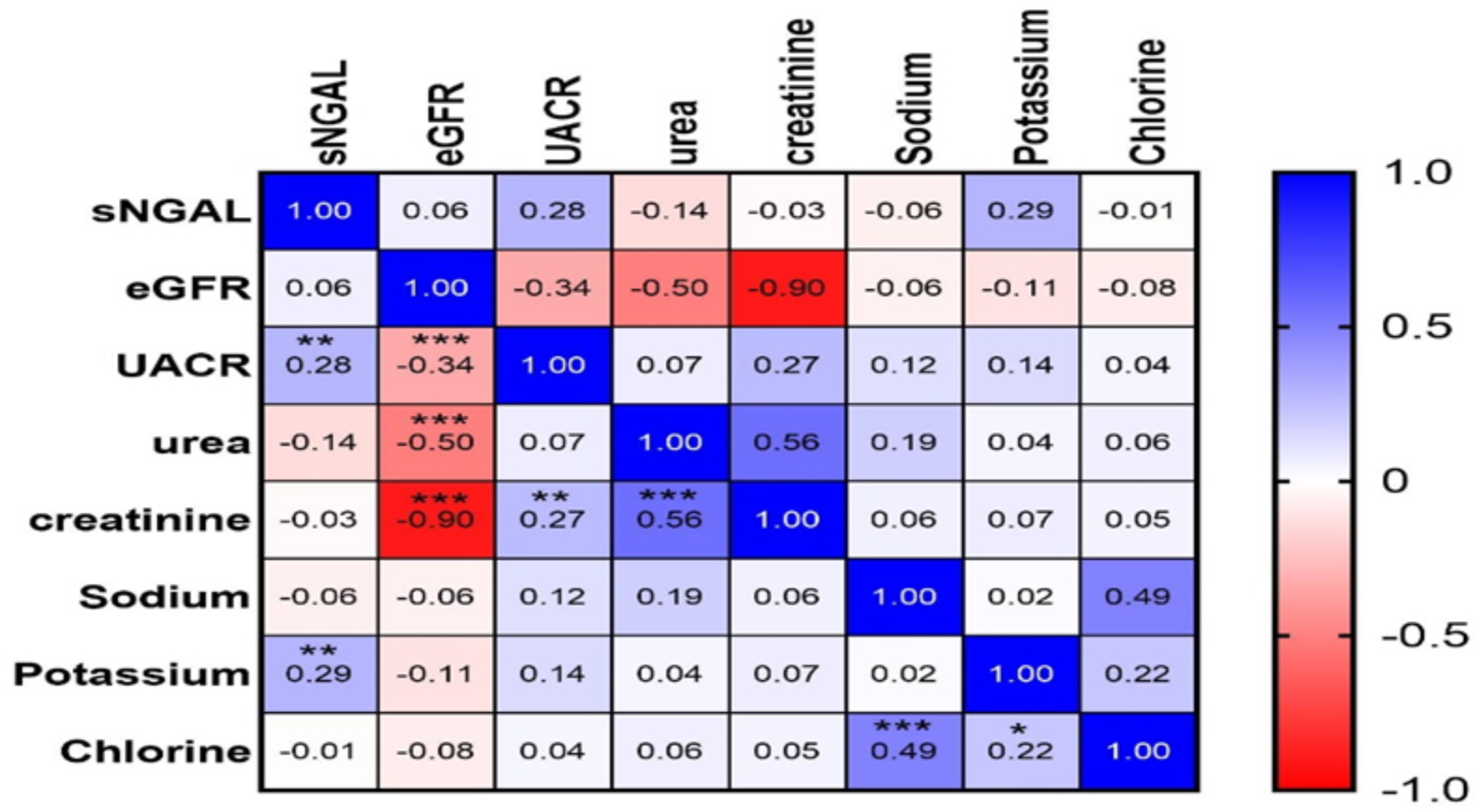
Heat map representing the correlation matrix between sNGAL, eGFR, UACR and the Renal function Tests (RFT); * Indicates the significance level of *p*< 0.05 of the correlation, **=*p*<0.005 and ***=*p*<0.0005. sNGAL; Neutrophil Gelatinase Associated Lipocalin eGFR; Estimated Glomerular Filtration Rate, UACR; Urine Albumin-to-creatinine ratio.

### Diagnostics performance of sNGAL, Creatinine, Urea and FAR in predicting nephropathy among SCD patients

Using the Receiver Operating Curve (ROC) analysis for predicting nephropathy among SCD patients, sNGAL as a marker at a cutoff of >5.72 significantly predicted nephropathy among SCD patients with high Area under the Curve (AUC=0.742, *p<0.0001*) and was very sensitive (91.2%) but less specific (51.4%). Also, Creatinine at a cutoff of >96.0 marginally predicted nephropathy among SCD patients significantly with AUC=0.618, *p=0.048* however, it was less sensitive (47.1%) but specific (81.4%). Urea and FAR predicted with an AUC=0.531 and AUC=0.462 respectively but were not statistically significant (*p>0.05*) (**Table 3** and **Figure 2**).

**Figure 2:**
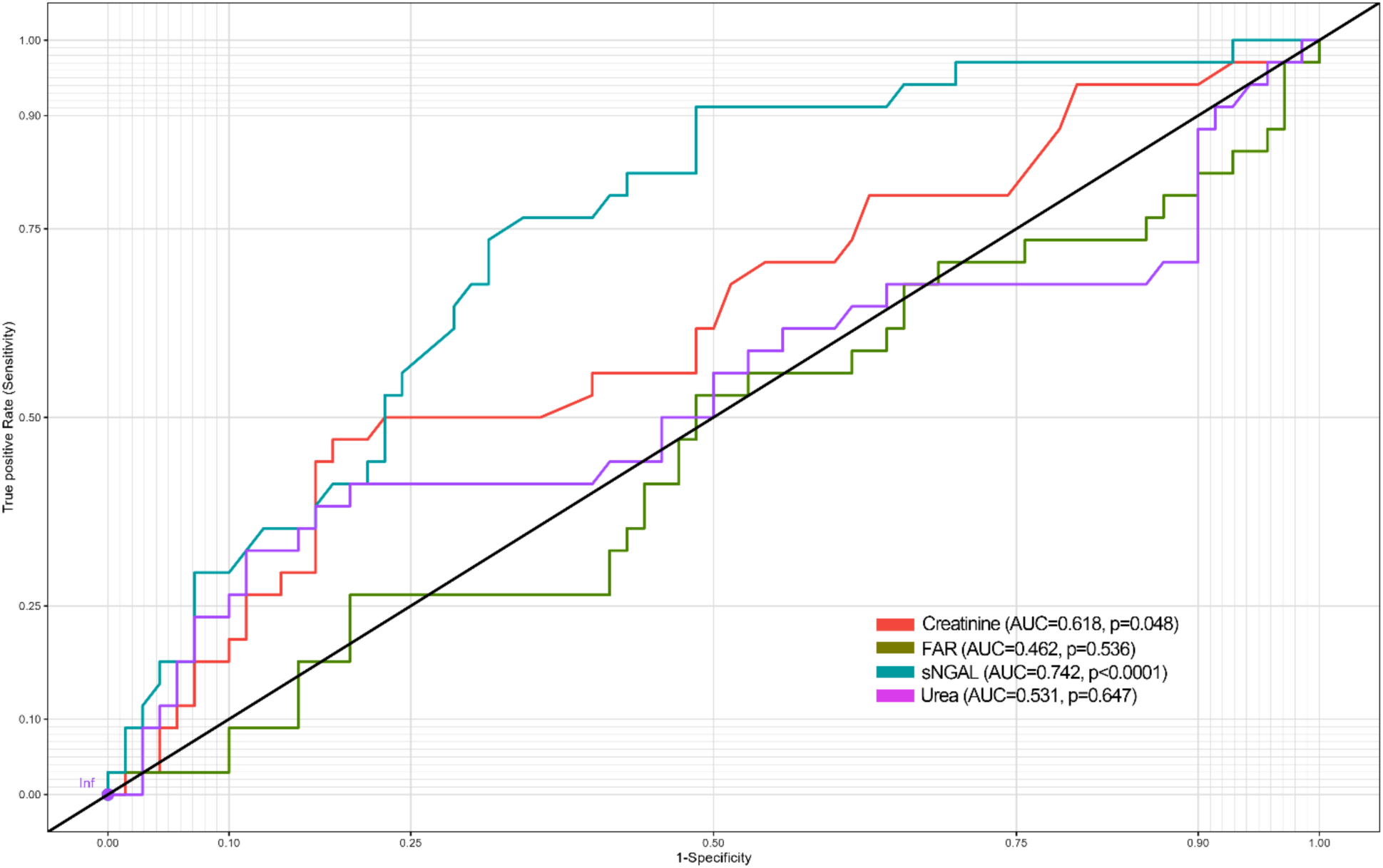
Receiver Operating Curve (ROC) for the various markers in predicting SCD nephropathy

**Table 3:** Diagnostic performance of sNGAL, Creatinine, Urea, and FAR, in predicting Nephropathy among SCD patients.

| Marker | Cut-off | Sensitivity (95% CI) | Specificity (95% CI) | PPV | NPV | LR+ | LR- | p-value |
| --- | --- | --- | --- | --- | --- | --- | --- | --- |
| sNGAL | >5.72 | 91.2 (76.1-97.6) | 51.4(40.0-62.7) | 47.7 | 92.3 | 1.88 | 0.17 | **<0.0001** |
| Creat | >96.0 | 47.1(31.5-63.2) | 81.4(70.6-88.9) | 55.2 | 76.0 | 2.534 | 0.65 | **0.048** |
| Urea | >3.06 | 41.2(26.4-57.8) | 80.0(69.0-87.8) | 50 | 73.7 | 2.059 | 0.735 | 0.649 |
| FAR | >0.12 | 26.5(14.5-43.4) | 80.0(69.0-87.8) | 39.1 | 69.1 | 1.324 | 0.919 | 0.536 |

Abbreviations: FAR; Fibrinogen-Albumin Ratio, Crea: Creatinine, sNGAL: serum Neutrophil Gelatinase – Associated Lipocalin, CI: Confidence interval, PPV: Positive predictive value, NPV: Negative predictive value, LR+: Positive likelihood ratio, LR–: Negative likelihood ratio

## Discussion

Renal involvement in SCD is demonstrated by the presence of crescent RBCs in the renal medulla, which cause ischemia, microinfarcts, reduced renal medullary blood flow, and renal papillary necrosis (14). Conventional renal markers remain normal until extensive renal damage hence the necessity for early diagnostic markers allows for implementation of preventive actions (15). This case-control study assessed NGAL and FAR as indicators for nephropathy among non-hospitalized SCD patients who were unaware of their renal status.

Most of the SCD patients were individuals with HbSS variant followed by HbSC variant which agrees with a research mastered by Ohene Frimpong *et al.*, (2008) and Ephraim *et al*., (2015) from Ghana (16). Ko *et al*., (2017) (17) reported that high protein intake can cause injury to the glomerular structure resulting in CKD which favors the high protein intake association with nephropathy in SCD subjects. However, restriction of protein intake is not recommended mainly due to the low energy state of SCD patients especially those with HbSS which may also explain the high protein intake in sickle cell nephropathy.

Prevalence of nephropathy among adult steady state SCD participants was 32.7% as indicated in 2024 KDIGO classification(12). This is above a prevalence of 17.1% nephropathy based on microalbuminuria found by Marouf et al., (2021) in Kuwait. However, this is below the prevalence indicated in a study by Ephraim *et al.,* (2015) that found that 68.4% of adult SCD patients had CKD based on reduced eGFR in Ghana. The variation in the prevalences might be due to the fact that eGFR only explore the function of the kidney whilst microalbuminuria determines kidney damage as indicated in KDIGO classification.

With respect to FBC parameters, there was lower haemoglobin and RBC levels in SCD nephropathy (SCN) participants compared to controls and SCD non-nephropathy (Non-SCN) participants. This confirms the significant association between haemoglobin, RBC and hematocrit levels and kidney disease as already indicated by various studies(18–22). We also reported significant diferrence between SCN, Non-SCN and healthy individuals with respect to total WBC levels and differentials in a decreasing order which tells the story that SCD is an inflammatory disease and get worse when there is an additional inflammatory disorder like nephropathy which agrees with several studies (5, 23, 24). Also, in a prospective study conducted by Aneke *et al.*, (2014) in Nigeria reported high levels of platelets in SCD participants with CKD which is in agreement with this present study (25). The pattern of WBC and platelets levels among participants justify the increase in inflammation in the SCN group resulting in high levels of NGAL, FAR and renal parameters. With respect to LFT parameters, ALT which is very specific to liver was apparently normal and similar in participant groups which rules out liver dysfunctyion in study participants. LFT markers seen in this study is in accordance with a research done by Kotila *et al*., (2005) who observed minimal elevation of transaminases in steady state SCD patients (26). As far as the kidney biomarkers are concern, this study reported markely increased levels of creatinine and urea which conforms to a research done by Ephraim *et al*., (2015) in Ghana. There was a minimal increased levels of potassium in the SCN compared to the controls which agrees with findings from Alhwiesh *et al.*, (2015).

NGAL was significantly higher in the SCN group as compared to the healthy controls. Marouf *et al.,* (2020) and Atere *et al*., (2018) found increased levels of NGAL in steady state SCD pateints and its relationship with renal abnormailities as comapred to healthy individuals which is consistent with this present study. Fibrinogen-to-albumin ratio was significantly higher in the SCN as comapared to Non-SCN and healthy controls(27). Increased fibrinogen and hypoalbuminuria are biomarkers of inflammation, this was evident by a study conducted by Luyendyk *et al*., (2019) and Zhang *et al*., (2019), who also found their correlation with renal prognosis which agree with this study. This study found a significant positive correlation between NGAL and UACR which is in line with a study performed by Marouf *et al.,* (2021) who found a positive non-significant correlation between plasma NGAL and UACR but a positive significant correlation between urine NGAL and UACR. This observation could be attributed to the state of pateints in that they recruited both steady and VOC SCD and they were not grouped in the correlation analysis. There was also a negative statistically significant correlation between eGFR and UACR. Also, there was a significant strong negative correlation between eGFR and a mild significant negative relationship existed between urea and eGFR. These observations are comparable to findings from (4, 5, 15, 28, 29). The optimal threshold at which NGAL level indicated impairment of the of kidney was 5.72ug/l with a sensitivity of 91.2% and specificity of 51.4%. Comaparing the Area under the curves NGAL significantly performed better than the conventional markers (urea and creatinine) with higher sensitivity (91.2%) than creatinine (47.1%) and that of urea (41.2%). However, creatinine was highly specific but the AUC value was lower than that of NGAL, unlike urea that had non-significant specificity and lower AUC value. This tells an interesting story that once there is kidney impairment in SCD individuals in steady state, NGAL would be able to detect it and can be used as a predictive marker for nephropathy. These observations are comparable to a research conducted by Atere *et al*., (2018) in Nigeria who comapared the AUC of NGAL and conventional markers (Urea and Creatinine) with NGAL having a superior AUC than urea and creatinine in both steady and VOC states. However, Marouf and associates indicated that urine NGAL performs better than plasma NGAL in predicting nephropathy in SCD in VOC state but could not find that of SCD in steady state. Various studies have also compared NGAL levels in other non-SCD nephropathy studies indicating that NGAL was a better marker for nephropathy even in non-SCD patients which should be well looked at (29–32). Fibrinogen-to-albumin ratio performed poorly in the ROC analysis with a non-significant low sensitivity and specificity which indicates that FAR levels could not be used to predict nephropathy in SCD subjects as seen in a cancer study by Hwang *et al*., (2017) which is inconsistent with this study, though FAR could be used to track nephropathy treatment due to the higher levels in SCD nephropathy group in this study.

Despite the novel findings, this study had limitations worth mentioning. The study recruited only steady state SCD patients and thus not entirely a true reflection of all SCD adult patients. Also, most of the SCD participants were young adults and may not be a reflective of old SCD patients. Further studies should recruit both steady state and VOC state adult SCD patients with equal age distribution. Urine albumin-to-creatinine ratio should be added to estimated glomerular filtration rate as suggested by KDIGO to confirm chronic kidney disease and thus classify sickle cell nephropathy. Further studies should be done to assess relationship between serum NGAL and urine NGAL and how they predict sickle cell nephropathy.

## Conclusion

In conclusion, this is the first attempt to investigate NGAL and FAR as indicators of nephropathy among adult steady state SCD patients in Ghana.The study demonstrated a higher NGAL and FAR levels among SCD with nephropathy as compared to apparently healthy individuals. The study demonstrated a significant correlation between Neutrophil Gelatinase- Associated Lipocalin and Albumin-to-Creatinine Ratio (UACR). Finally, NGAL as independent marker, is an early predictor of kidney disease as compared to urea and creatinine. Fibrinogen-to-albumin ratio can be used to track nephropathy treatment in steady state sickle cell disease patients since it is elevated in those with sickle cell nephropathy compared to healthy individuals.

## Data Availability

All data produced in the present study are available upon reasonable request to the authors

## Acknowledgements

We are grateful for the immense contributions of the staff of Komfo Anokye Teaching Hospital, Suntreso Government Hospital and Kumasi South Hospital for their warm reception, not forgetting our participants. Special thanks to the serology departrment of Komfo Anokye Teaching Hospital on their support in storage of samples.

## Authors’ contributions

**Conceptualization:** Stephen Twumasi, Lilian Antwi Boateng, Enoch Odame Anto, Benedict Sackey, Allwell Adofo Ayirebi

**Data curation**: Stephen Twumasi, Lesle Osei, Angela Opoku

**Formal analysis**: Stephen Twumasi, Richard Owusu Ansah, Godfred Yawson Scott, Alfred Effah **Investigation**: Stephen Twumasi, Lilian Antwi Boateng, Enoch Odame Anto, Angela Opoku, Leslie Osei, Vivian Paintsil, Joyce Duku, Emmanuel Asafo Adjei

**Methodology** : Stephen Twumasi, Lilian Antwi Boateng, Enoch Odame Anto

**Project administration**: Stephen Twumasi, Lilian Antwi Boateng, Enoch Odame Anto

**Resources:** Stephen Twumasi, Angela Opoku

**Software**: Stephen Twumasi,Richard Owusu Ansah, Godfred Yawson Scott, Alfred Effah

**Supervision**: Stephen Twumasi, Lilian Antwi Boateng, Christian Obirikorang, Enoch Odame Anto

**Validation**: Stephen Twumasi, Lilian Antwi Boateng & Enoch Odame Anto

**Visualization**: Stephen Twumasi, Lilian Antwi Boateng & Enoch Odame Anto

**Writing-original drtaft**: Stephen Twumasi, Lilian Antwi Boateng, Enoch Odame Anto, Richard Kobina Dadzie Ephraim, Christian Obirikorang, Benedict Sackey, Allwell Adofo Ayerebi **Writing-review & editing:** Stephen Twumasi, Enoch Odame Anto, Benedict Sackey, Christian Obirikorang, Alfred Effah, Richard Kobina Dadzie Ephraim, Allwell Adofo Ayirebi, Godfred Yawson Scott, Richard Owusu Ansah, Angela Opoku, Leslie Osei, Vivian Paintsil, Joyce Duku, Emmanuel Asafo Adjei, Lilian Antwi Boateng

## Availability of data

Data and materials for study are available upon request from the corresponding authors.

## Declarations

The study was approved by the Committee on Human Research and Publication Ethics of the Kwame Nkrumah University of Science and Technology (CHRPE/AP/1037/23) and Komfo Anokye Teaching Hospital (KATH IRB/AP/175/23). Written consent of individual participants was sought after the aims, benefits, risk and right of withdrawal at any time from the study were well explained to the study participants in English and in the local dialect (mostly Twi) and their consent was obtained.

## Consent for publication

Not applicable.

